# Pediatric Nemaline Myopathy: A systematic review using individual patient data

**DOI:** 10.1101/2022.01.31.22270131

**Authors:** Briana Christophers, Michael A. Lopez, Vandana A. Gupta, Mary Baylies

**Affiliations:** Weill Cornell/Rockefeller/Sloan Kettering Tri-Institutional MD-PhD Program, New York, NY; University of Alabama Birmingham, Birmingham, AL; Brigham and Women’s Hospital and Harvard Medical School, Boston, MA; Sloan Kettering Institute, Memorial Sloan Kettering Cancer Center, New York, NY

## Abstract

**Context:** Nemaline myopathy (NM) is a skeletal muscle disease that affects 1 in 50,000 live births.

**Objective:** Develop a narrative synthesis of the findings of a systematic review of the latest case descriptions of patients with nemaline myopathy.

**Data Sources:** A systematic search of MEDLINE, Embase, CINAHL, Web of Science, and Scopus was performed using Preferred Reporting Items for Systematic Reviews and Meta-analyses (PRISMA) guidelines using the keywords pediatric, child, nemaline myopathy, nemaline rod, rod myopathy.

**Study Selection:** Case studies focused on pediatric NM and published in English between January 1, 2010 and December 31, 2020 in order to represent the most recent findings.

**Data Extraction:** Information was collected about the age of first signs, earliest presenting neuromuscular signs and symptoms, systems affected, progression, death, pathological description, and genetic changes.

**Results:** Of a total of 385 records, 55 case reports or series were reviewed, covering 101 pediatric patients from 23 countries. We review varying presentations in children ranging in severity despite being caused by the same mutation, in addition to current and future clinical considerations relevant to the care of patients with nemaline myopathy.

**Limitations:** The results are limited by findings shared in the literature, which varied in their detail and sometimes lacked histological or genetic results. Trends and correlations may be altered by information available.

**Conclusions:** This review synthesizes genetic, histopathologic, and disease presentation findings from pediatric nemaline myopathy case reports. These data strengthen our understanding of the wide spectrum of disease seen in nemaline myopathy. Future studies are needed to identify the underlying molecular mechanism of pathology, to improve diagnostics, and to develop better methods to improve quality of life for these patients.

## Introduction

Nemaline myopathy (NM) is a primary skeletal muscle disease and histopathologic diagnosis with variable clinical presentation and genetic causes. It has an estimated incidence of 1 in 50,000 live births. ^1^ The disorder was first described in 1963 in the case of a child with hypotonia and “microgranules” in a muscle biopsy. ^2,3^ Clinico-histopathological diagnosis of several congenital myopathies includes the detection of such aggregates, distinguished as cores, electron-dense rods, or central nuclei. ^4^ Rods detected in the sarcoplasm or inside the nucleus leads to a diagnosis of NM. These rods are often seen emanating from the muscle Z-disc, the anchor point of the sarcomere unit (Figure 1). Sarcomeric proteins have been shown to make up a portion of these aggregates. ^5,6^ Additionally, individual muscle fibers may be affected by either atrophy or hypertrophy. ^7,8^

**Figure 1.**
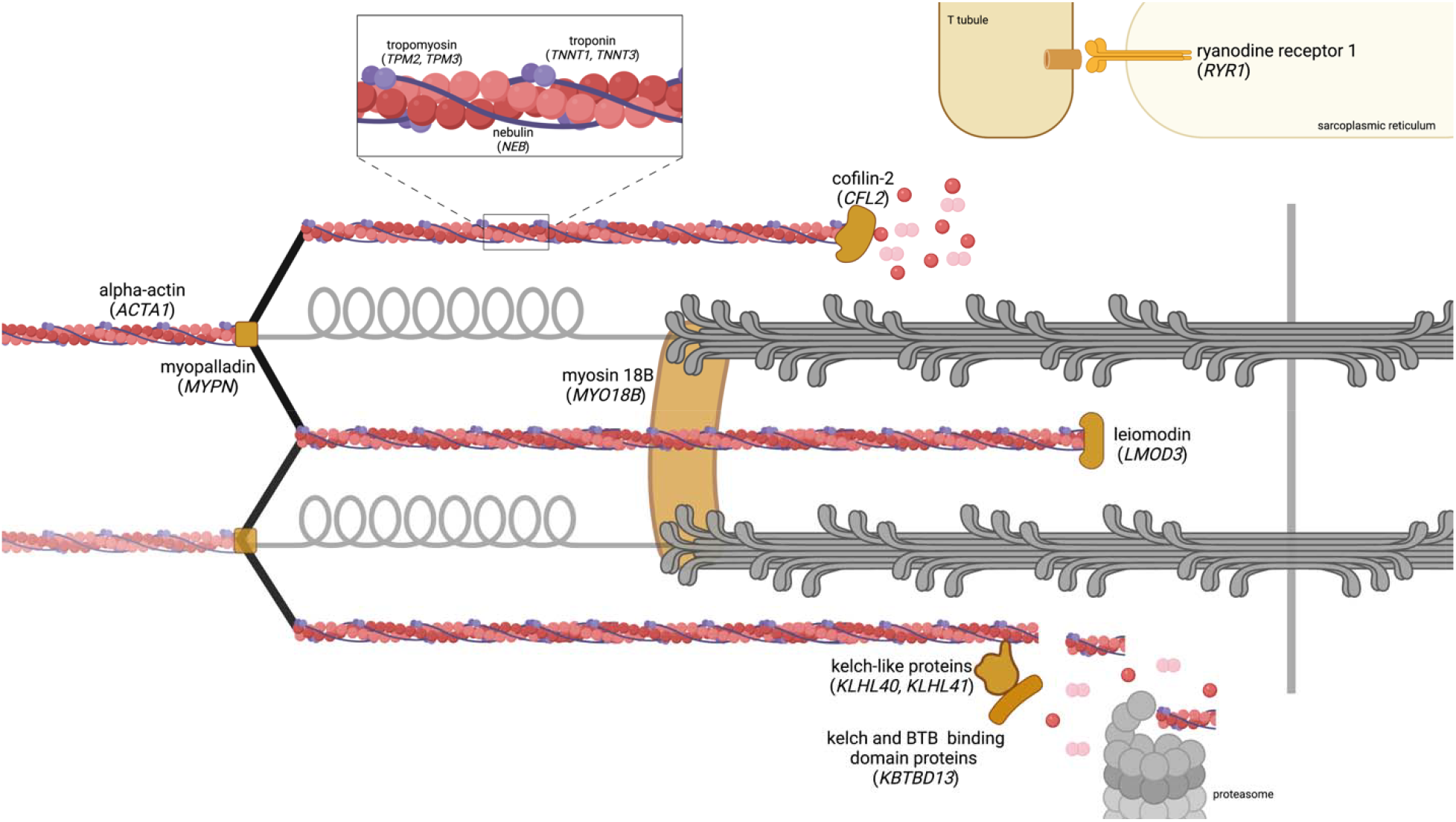
Sarcomere-related proteins causative of nemaline myopathy. Those genes designated with an asterisk (*) correspond to genes sequenced in the case studies reviewed presently. Created with BioRender.com.

To date, pathogenic variants in 12 genes have been identified as causative of NM (Table 1). ^9^ Several of the implicated proteins localize to the thin filament of the sarcomere: skeletal alpha-actin 1 (ACTA1), cofilin (CFL2), leiomodin 3 (LMOD3), nebulin (NEB1), and kelch repeat and BTB domain containing protein 13 (KBTBD13). ^10–15^ Mutations have also been found in Kelch-like family member proteins (KLHL40, KLHL41) that regulate the protein turn-over of thin filaments. ^16^ Additional structural or regulatory components of the sarcomere may be affected as shown by mutations in genes encoding myosin 18B (MYO18B), myopalladin (MYPN), ryanodine receptor 1 (RYR1), slow skeletal muscle troponin T (TNNT1), fast skeletal muscle troponin T (TNNT3), and slow muscle alpha-tropomyosin (TPM2, TPM3). ^17–21^ As far as the functions of the proteins, skeletal alpha-actin is the main component of the actin filament, which is structurally supported by nebulin, troponin, tropomyosin, and myopalladin. ^10,13,17,19,21^ Leiomodin is an actin nucleator, while cofilin is an actin-severing protein involved in maintaining the balance between filamentous and monomeric actin. ^11,12^ Meanwhile, the KBTBD13 adaptor protein works with KLHL40 and KLHL41 to regulate thin filament stability with the ubiquitin-proteasome system. ^15,16^

**Table 1.** Genetics of pediatric nemaline myopathy case reports (2010-2020). Genetic changes described in the cases reviewed presently, including number of cases by gene symbol, genetic and protein changes, and a description of protein function.

| Gene Symbol | Cases | Protein Function | Genetic and Protein Changes | References |
| --- | --- | --- | --- | --- |
| *ACTA1* | 22 | principal actin isoform of skeletal muscle thin filament | c.283C>A, p.Asn94Lys<br>c.350A>G, p.Asn117Ser<br>c.356G>A, p.Glu85Lys<br>c.407T>C, p.Val136Ala<br>c.430C>T, p.Leu144Phe<br>c.448A>G, p.Thr150Ala<br>c.455G>C, p.Gly152Ala<br>c.478G>A, p.Gly160Ser<br>c.487C>G, p.His163Asp<br>c.557A>G, p.Asp186Gly<br>c.611C>T, p.Thr204Ile<br>c.760A>C, p.Asn254His<br>c.801G>C, p.Gly270Arg<br>c.868G>A, p.Asp290Asn<br>c.871A>T, p.Tyr592HisfsX17<br>c.911delG, p.Gly304AlafsX24<br>c.1049C>T, p.Ser350Leu<br>c.1074G>T, p.Trp358Cys<br>c.1075A>C, p.Ile359Leu<br>c.1127G > C, p.Cys376Ser | Saito 2011<br>Yang 2016<br>Lehtokari 2018<br>Ennis 2015<br>Saito 2011<br>Miyatake 2014<br>Ravenscroft 2011<br>Yokoi 2019<br>Moreno 2017<br>Levesque 2013<br>Moreno 2017<br>Pula 2020<br>Saito 2011<br>Seidahmed 2016<br>Moureau-Le Lan 2018<br>Friedman 2014<br>Moreno 2017<br>Gatayama 2013<br>Yeşilbaş 2019<br>Waisayarat 2015 |
| *CFL2* | 6 | actin-severing and -binding protein | c.19G>A, p.Val7Met<br>c.100_103delAAAG, p.Lys34Glnfs*6<br>c.235G>T, p.Asp79Tyr<br>c.256G>C, p.Asp86His<br>c.281delC, p.Ser94LeufsTer6 | Ockeloen 2012<br>Ong 2014<br>Fattori 2018<br>Fattori 2018<br>Fattori 2018 |
| *KLHL4*<br>*0* | 10 | binding partner of E3 ligase cullin 3 | c.604delG, p.Ala202Argfs*56<br>c.1405G>T, p.Gly469Cys<br>c.1516A>C p.Thr506Pro<br>c.1327G>A, p.Gly433Ser<br>c.1498C>T, p.Arg500Cys<br>c.1513G>C, p.Ala505Pro<br>chr3:42727712G>A, p.Trp201Ter | Natera-de Benito 2016<br>Kawase 2015<br>Yeung 2020<br>Yeung 2020<br>Seferian 2016<br>Natera-de Benito 2016<br>Avasthi 2019 |
| *LMOD3* | 5 | actin nucleator | c.366delG, p.Lys122AsnFs*6<br>c.882dupA, p.Asp295Argfs*2 | Marguet 2020<br>Michael 2018 |
|  |  |  | c.1069G>T, p.Glu357*<br>c.1628G>T, p.Arg543Leu<br>p.Glu121ArgfsTer5<br>p.L245del | Michael 2018<br>Marguet 2020<br>Berkenstadt 2018<br>Berkenstadt 2018 |
| **MYO18B** | 2 | unconventional myosin, function not well characterized | c.6496G>T; p.Glu2166* | Malfatti 2015 |
| **NEB1** | 24 | structural component of sarcomere, binds actin | c.300dup, p.Tyr101fs*5 int49<br>c.3458+1G>A<br>c.1825dup, p.Pro541Profs*2<br>c.2414 + 5G>A<br>c.5060□G>A, p.W1687X<br>c.5343 + 5G>A,<br>p.Arg1747_Thr1778del<br>c.5574C□>□G, p.Tyr1858Stop<br>c.6496-G>A, p.2166_2234del<br>c.7432 + 1916_7535 + 372del,<br>p.Arg2478_ Asp2512del<br>c.11086A>C, p.Thr3696Pro<br>c.13066delT, p.Tyr4356Thrfs*8<br>ex110<br>c.17535G > A, p.Glu5845Glu<br>c.17779_17780delTA,<br>p.Tyr5927HisfsX17<br>c.18676C>T, p.Gln6226*<br>c.19101 □+□5G□>□A,<br>p.Leu6333_Glu6367del<br>c.20928G>T; p.Gly6976Gly<br>c.21076CC>T, p.Arg7026Ter<br>c.21796_21810delinsT,<br>p.Pro7266fs*30<br>c.22273del, p.Val7425Serfs49*<br>c.22591-3C>G, p.7531Val_<br>Ser7564del<br>c.2310+5G>A,<br>c.17779_17780delTA,<br>p.His738_Asp770del<br>c.23420_23421del,<br>p.Arg7807Serfs*16<br>c.24250_24253dupGTCA,<br>p.T8085fsX8100<br>c.24269del, p.Arg8090fs*54<br>c.24372_24375dupAAGA,<br>p.Val8126fs<br>c.24440_24441insGTCA, | Malfatti 2014<br>Kapoor 2012<br>Lehtokari 2011<br>Lehtokari 2011<br>Kiiski 2015<br>Malfatti 2014<br>Malfatti 2014<br>Malfatti 2014<br>Malfatti 2014<br>Moureau-Le Lan 2018<br>Malfatti 2014<br>Malfatti 2014<br>Moureau-Le Lan 2018<br>Malfatti 2014<br>Malfatti 2014<br>Malfatti 2014<br>Moureau-Le Lan 2018<br>Moureau-Le Lan 2018<br>Moureau-Le Lan 2018<br>Malfatti 2014<br>Malfatti 2014<br>Moureau-Le Lan 2018<br>Malfatti 2014<br>Gajda 2015<br>Malfatti 2014<br>Scoto 2013<br>Malfatti 2014<br>Gajda 2015<br>Malfatti 2014<br>Malfatti 2014<br>Malfatti 2014 |
|  |  |  | p.Pro8148Serfs*15<br>c.24527_24528delCT,<br>p.P8176fsX8179<br>c.24579G>A, p.Ser8193Ser ex119<br>c.24686_24687del,<br>p.Glu8229Glu fs*18<br>c.24735_24736del(AG),<br>p.Arg8245fs*1<br>c.163689G>T, GAG>UGA<br>hg19<br>chr1:g.(154,156,325_154,156,028)_(<br>154,173,059_154,177,712) | Kapoor 2012<br>Kiiski 2015 |
| ***RYR1*** | 2 | intracellular calcium channels at the sarcoplasmic reticulum | c.4455-4G>A<br>c.4718C>T, p.Pro1573Leu<br>c.7585G>A, p.Asp2529Asn | Tiberi 2020<br>Kondo 2012<br>Kondo 2012 |
| ***TNNT1*** | 15 | muscle contraction regulator | c.309+1G>A<br>c.323C>G, p.Ser108X<br>c.574_577delinsTAGTGCTGT<br>c.661G>T, p.Glu221X<br>arr[GRCh37]<br>19q13.42(55652193_55663445)x0 | van der Pol 2014<br>Marra 2015<br>Abdulhaq 2015<br>D'Amico 2019<br>Streff 2019 |
| ***TPM2*** | 2 | muscle contraction regulator | c.415_417delGAG, p.E139del | Citirak 2014 |
| **Unkno**<br>**wn** | 13 |  |  |  |

NM symptoms may present at any point during fetal development to adulthood. Typically, NM presents similarly to other congenital myopathies: hypotonia, muscle weakness, and decreased or absent myostatic stretch reflexes. Defects are observed in the musculoskeletal system, such as fractures, kyphoscoliosis, joint contractures, skull enlargement, pectum excavatum, and pes planus are observed. Creatine kinase levels are normal, although this is also the case in other myopathies. Many patients are born with characteristic myopathic facies affecting mainly the mouth resulting in a high-arched palate and drooling; occasionally patients present with ophthalmoplegia which is rarely seen in other musculoskeletal conditions. Complications in the respiratory system are common. ^22^ In rare cases, cardiac and renal systems may also be affected secondary to the musculoskeletal defects. ^23–25^ The differential diagnosis for patients with severe hypotonia and bulbar weakness is broad, but at birth includes myotonic dystrophy type 1, congenital myasthenic syndromes, spinal muscular atrophy type 0/1, mitochondrial myopathies, and Pompe disease. ^26,27^

The goal of this systematic review was to synthesize latest published findings in pediatric patients with NM as an initial step toward describing genotype-phenotype descriptions. Our examination of case reports published between 2010 and 2020 details novel genetic findings, newly described clinical presentations, and clinical considerations for the care of pediatric NM patients.

## Methods

A systematic search of Pubmed, Embase, CINAHL, Web of Science, and Scopus was performed using Preferred Reporting Items for Systematic Reviews and Meta-analyses (PRISMA) guidelines. ^28^ The search strategy used the keywords (pediatric* OR child*) AND (nemaline myopathy OR nemaline rod OR rod myopathy). This protocol was not registered. Included case series or studies were published between January 1, 2010 and December 31, 2020; originally published in English; had pediatric patients (<21 years old); and focused on NM as the main topic. The time period selected limited our search to publications with the most recent peer-reviewed findings on pediatric NM. Reviews, meta-analyses, book chapters, and abstracts were excluded. Selected references were exported to Sciwheel at which point all duplicates were removed.^29^ Titles were read for concordance with inclusion criteria as a screen, after which abstracts were read to select for eligible articles. Full text of all eligible articles was read, and any that did not fulfill the inclusion criteria were removed from consideration. These full text articles fulfilled the CARE guidelines for case reports.^30^ Data was extracted and tabulated about age of first signs, earliest presenting neuromuscular signs and symptoms, accompanying body systems affected, progression, death, pathological description, genetic changes, and consanguinity. These data were synthesized with attention to the recommendations of “Methodological quality and synthesis of case series and case reports.” ^31^ Extracted data is available upon request.

## Results

The database search yielded a total of 385 records (Figure 2). Of the 69 eligible articles, 55 case reports or series of 101 pediatric patients from 23 countries were selected for this review (Supplement). By genetic mutation, reports implicated *ACTA1, KLHL40, NEB*, and *TNNT1*, in addition to other genes with a minor contribution (Table 1).

**Figure 2.**
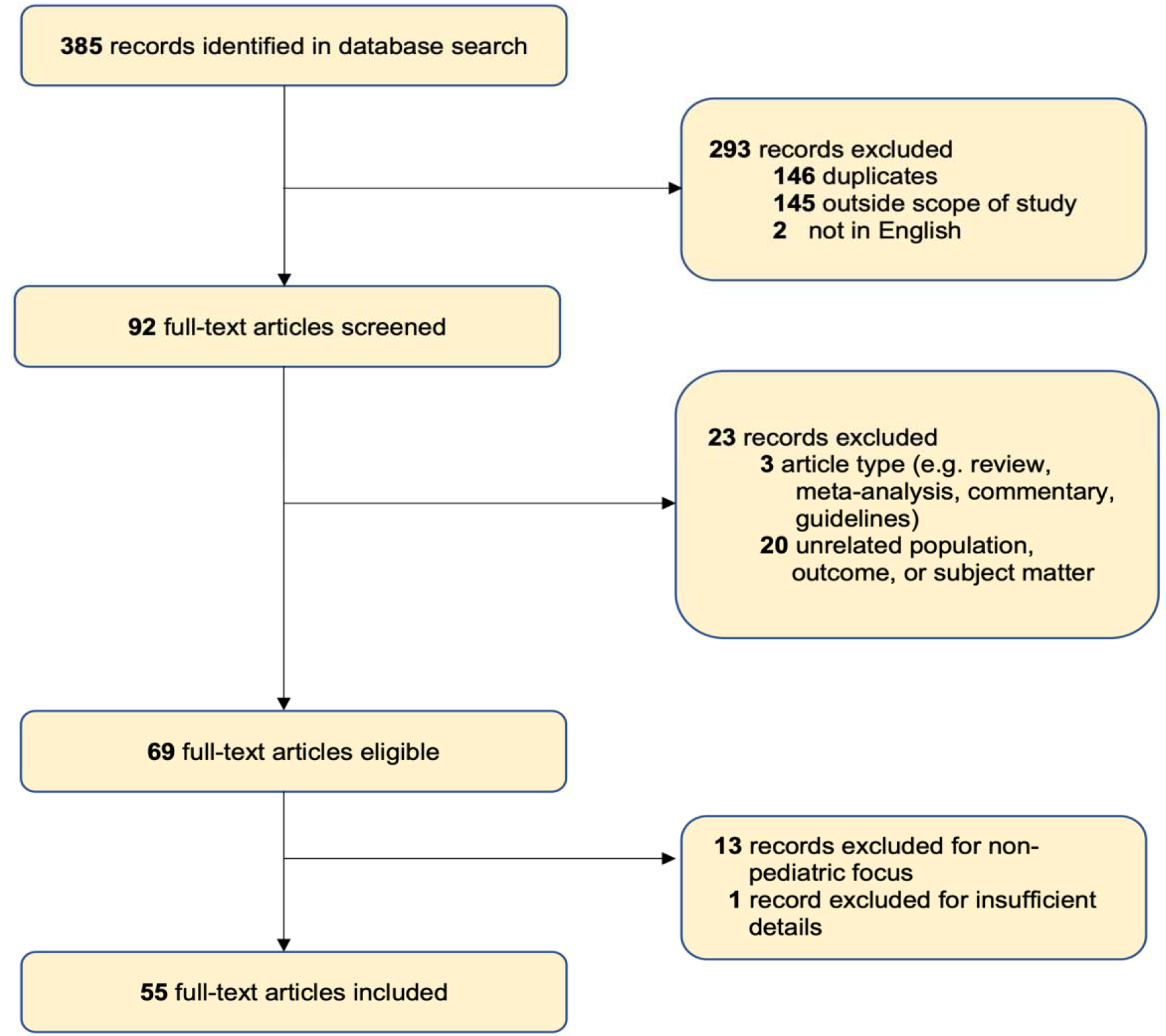
Study Selection Process using PRISMA Flow Diagram.

### Genetics

Improved clinical availability of next generation sequencing has increased identification of novel mutations associated with pediatric NM, especially exome sequencing. ^32^ The 88 cases covered by this review with available genetic information implicate several variants as causative of NM: 24% deletions and 6% duplications. These resulted in 25% missense mutations and 9% frameshift mutations. These changes were predicted to cause protein truncations in 19% of the cases. NM mutations were found to be *de novo* in 11% of cases. Further examination of the sequencing data revealed that 30% of the patients were heterozygous for the mutated gene, 23% were homozygous, and 7% were compound heterozygous.

*NEB* and *ACTA1* have been identified as the most commonly mutated genes in NM. Of the pediatric cases examined for this review, 25% had mutations in *NEB* while 22% in *ACTA1*. Consistent with previous literature, many of the *NEB* mutations were frameshifts and present in compound heterozygous combinations. ^33^ 50% of the *ACTA1* mutations were missense and 45% were reported as *de novo* in the proband. For two patients, the *ACTA1* mutations were predicted to produce a truncated protein or affect the binding site for actin’s interactors. Three cases with *ACTA1* mutations had parents with gonadal mosaicism, which had not been previously reported, while one case showed somatic mosaicism. ^34–36^ Identifying mosaic mutations required additional in depth sequencing due to being very-low-grade.

New mutations were found in less common causative genes, such as *CFL2, KLHL40, RYR, TNNT1*, and *TPM3* (Table 1). New variants in *TNNT1* were documented in non-Amish populations. ^37–39^ van der Pol *et al*. predict that the combination of splice site mutations and a deletion in TNNT1 would produce short in-frame transcripts that would be targeted by nonsense-mediated decay. ^39^ One family harbored a novel four-base-pair deletion in *CFL2* likely causing a premature stop codon in the protein. ^40^ Three patients from two unrelated families described by Fattori *et al*. had mutations that may cause misfolding, lack of actin-binding, and protein degradation of cofilin 2. ^40,41^ In southern China, researchers identified a *KLHL40* founder mutation inherited both homozygous and heterozygous, which often leads to a truncation of the protein. ^42^ Several new mutations in tropomyosin genes, including *TPM3*, have also been identified. ^43,44^ *RYR* mutations have been associated with various myopathies, but rarely with NM. Massively parallel sequencing in one patient in this review brought to light a new mutation in *RYR1*, causing NM with ophthalmoplegia. ^45^

### Histopathologic features

Pathology information was available for 64% of the patients reviewed (Table 2). Of these, 75% confirmed presence of cytoplasmic nemaline rods or bodies. Curiously, one case report about a patient with an *LMOD3* mutation noted that the rods were surrounded by a filamentous halo. ^46^ Eight percent of patients had intranuclear rods; four of the five patients with intranuclear rods had a confirmed mutation in *ACTA1* with the fifth not having any information about genetic causes. Internalized nuclei (found in 9%) were seen in patients with different mutated genes, including *ACTA1, CFL2*, and *TNNT1*. ^38,41,44,47^

**Table 2.**
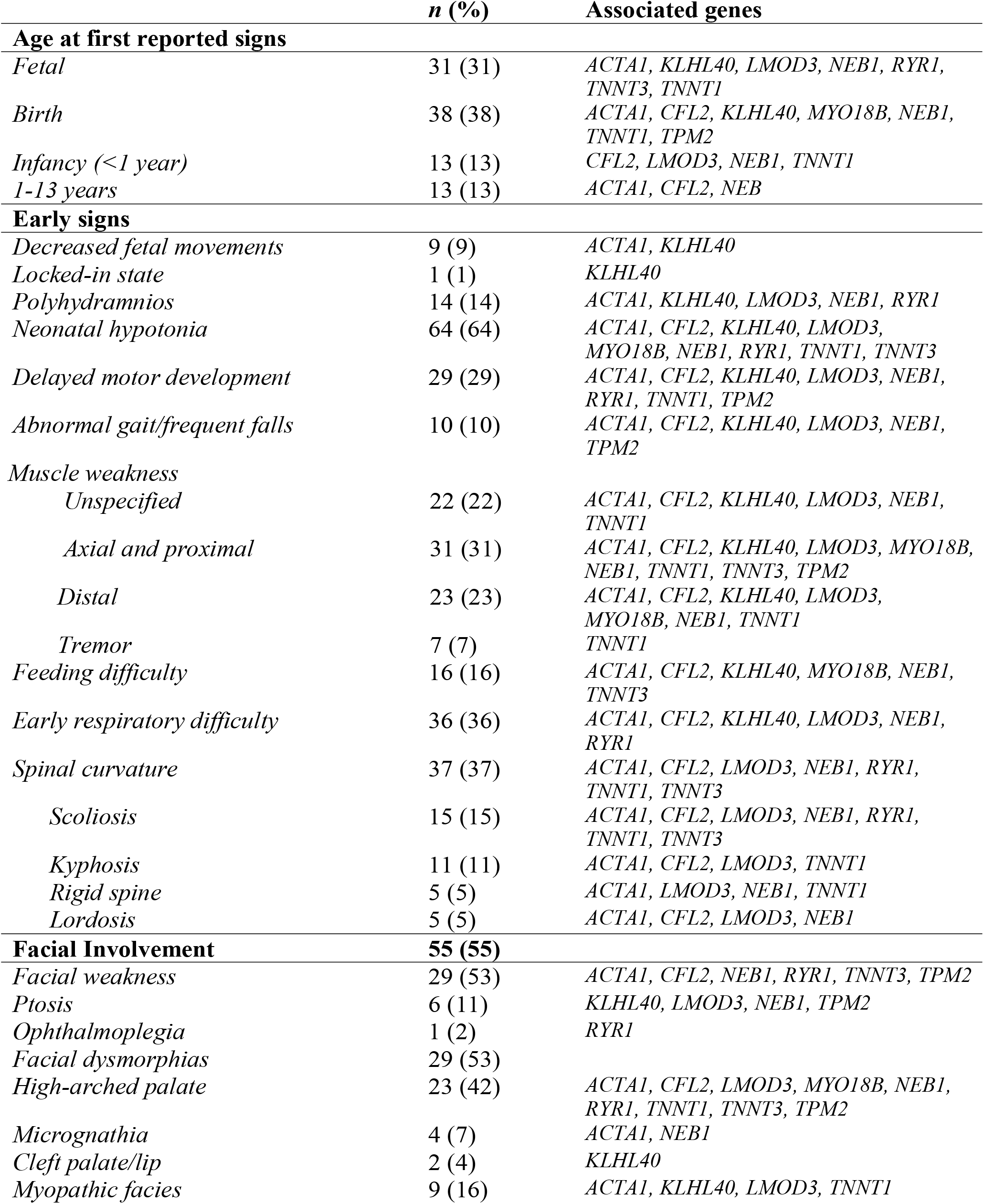
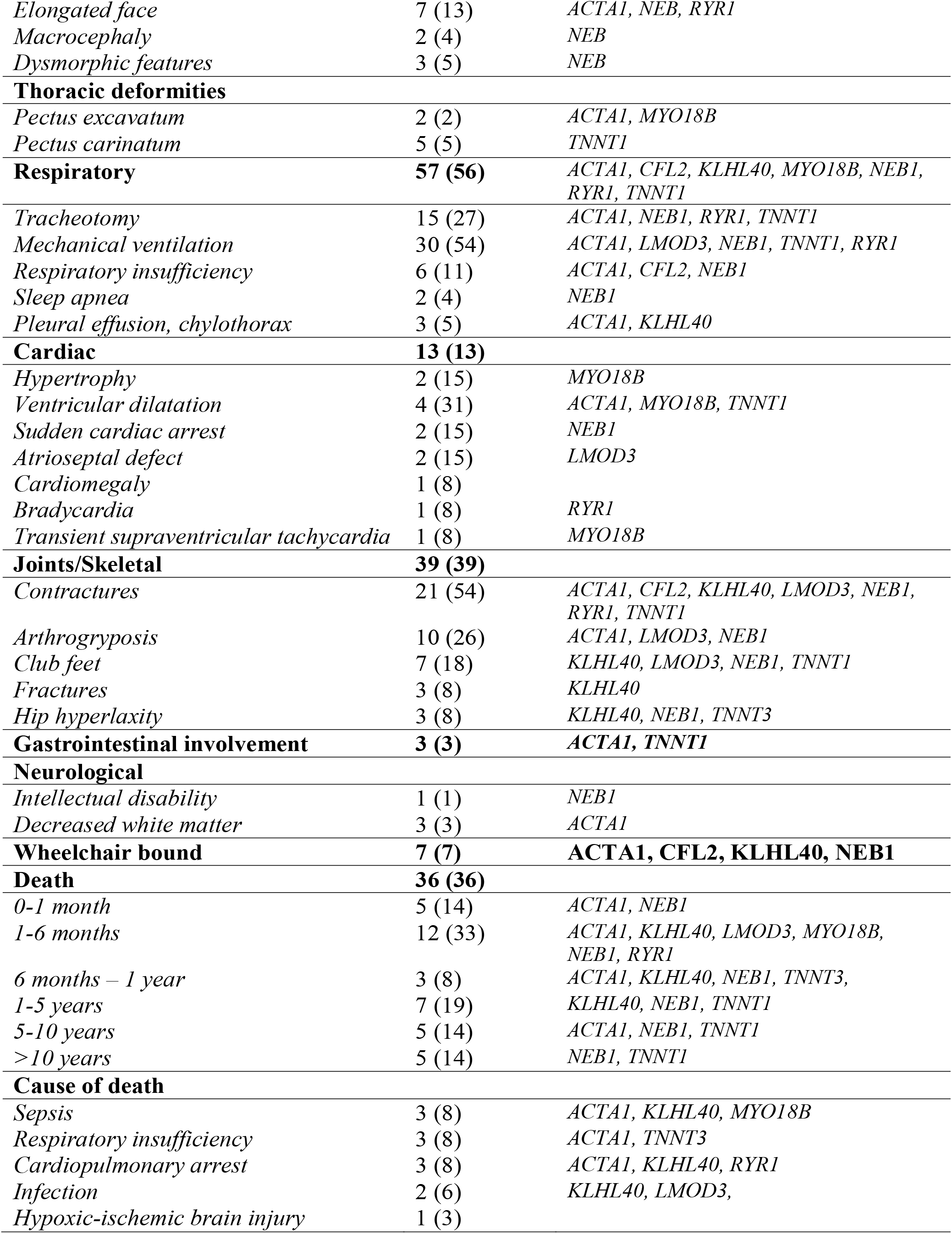
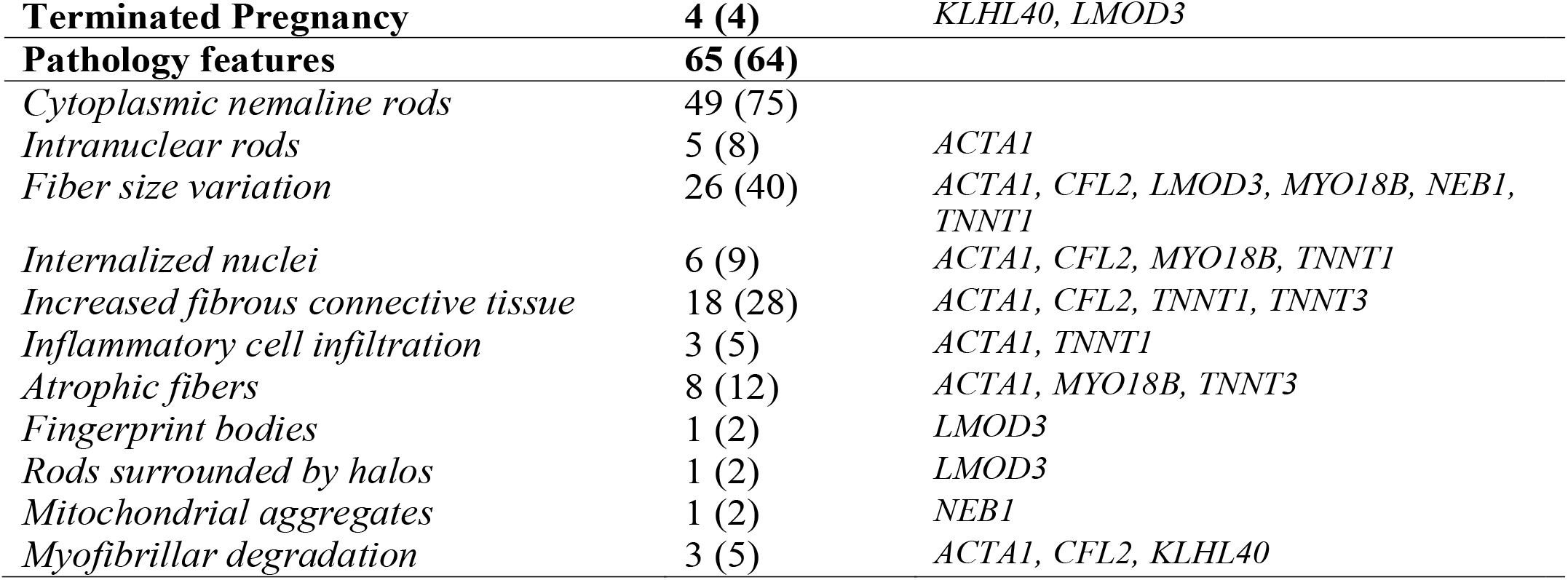
Main clinico-pathological features reported.

Other common features included fiber size variation (40%) and increased endomysial fibrous connective tissue (28%). Five percent of cases described some form of inflammatory cell infiltration. One study found that two patients had necrotic muscle fibers with T cells that stained positive for CD4 and negative to CD8. ^48^ Atrophic fibers were found in 12% of patients with no correlation to age of first symptoms or death at time of case report. Detailed analysis of patients with *NEB* mutations categorized them into three groups by time of symptom onset which correlated with differences on histology such as differences in the amount of sarcomeric dissociation, rod pattern and fiber type. ^49^

### Spectrum of Disease

The selected case reports described a variety of clinical presentations, some with symptoms in organ systems beyond the typical muscle weakness of NM (Table 2). The first reported signs often came as early as fetal development for 9% of cases with the report noting a decrease in fetal movements or polyhydramnios on antenatal ultrasound, typically during the second or third trimesters. The most common early sign of disease was neonatal hypotonia, seen in 64% of children, followed early respiratory difficulty (36%) and spinal curvature (37%). Over half of patients had respiratory complications throughout childhood, including respiratory failure as evidenced by placement of tracheostomy. Mutations in *MYO18B, TNNT1*, and *ACTA1* were seen in some cases that involved cardiomyopathy. ^17,50–52^ One patient presented early with left-sided hypertrophy causing pulmonary hypertension and right ventricular dilatation. ^17^ Dilated cardiomyopathy was identified in two patients with *ACTA1* mutations; this was paired with dyskinesia of the left ventricle in one child. ^34,51^

Muscle weakness presented differently across patients. Pattern of weakness was axial and proximal in 31% of cases, while distal involvement was reported for 23%. Facial weakness was noted for just under one-third of children, although a broad spectrum of facial dysmorphic patterns was also described. Some patients are born with significant craniofacial deformities, including cleft lip, atrophy of facial and masticator muscles, and jaw deformity, which can impact the patient’s ability to close their mouth. ^53^ Joint contractures and arthrogryposis were common, described in almost 40% of patients. Another possible presenting symptom is hyperextension of the neck, for which Tiberi *et. al*. recommend neurological examination after birth to check for possible muscle disorder such as NM. ^54^ One patient with decreased movement *in utero* was found to have very fragile ribs on postnatal chest radiograph. ^26^

The majority of patients exhibiting early lethality (36% of patients were deceased) were diseased within the first year of life; causes of death included sepsis, respiratory insufficiency, cardiopulmonary arrest, infection, or hypoxic-ischemic brain injury.

Patients with NM typically have normal intelligence, but Saito *et al*. documented patients with *ACTA1* dominant missense mutations with delays on word comprehension testing that “may result from abnormal development of the central nervous system, not only from hypoxic events or limited social experiences.” ^55^ All three patients had decreased white matter volume, enlargement of the lateral ventricles, and frontal lobe hypoplasia.

Clinical severity varied significantly, even amongst patients with mutations in the same gene. Some with *ACTA1* mutations were reported to have mild phenotypes, while one patient had dysautonomia in addition to myopathic symptoms. ^56–58^ Anticipation was seen in a family harboring an *ACTA1* missense mutation, where the mother had some weakness: her child had myopathic facies, high-arched palate, lumbar hyperlordosis, and delayed motor milestones. ^59^ One child with an *ACTA1* mutation of Thai ancestry had primary pulmonary lymphangiectasia on histology, in addition to pleural adhesions and a choroid plexus papilloma causing communicating hydrocephalus. ^47^ For *KLHL40* mutations, some patients had moderate presentation while others experienced fetal akinesia or a total locked-in state due to the severity of their disease. ^60–62^ *LMOD3* mutations also could present prenatally as decreased fetal movements, polyhydramnios, and arthrogryposis, or later in childhood, with a milder phenotype or even disease progression. ^12,46,63^ Mutations in *TNNT1* were reported in patients of varied ethnic backgrounds, including a case series of Amish patients presenting with the prototypical progressive muscle weakness, contractures, and tremors; of Italian siblings with severe failure to thrive and rigid spine; and of a Palestinian cohort which showed signs of transient tremors, progressive spinal rigidity, and limb contractures. ^64–66^

### Clinical Care Considerations

Care for patients with nemaline myopathy should follow standard multi-disciplinary supportive care as reviewed in the Consensus Statement on Standard of Care for Congenital Myopathies. ^67^ Biopsy is critical to the diagnosis of NM; however selection of the biopsy location may introduce sampling error. A potential advance was discussed by one paper that showed the potential of using imaging—magnetic resonance imaging, in particular—to guide the decision of what muscle to biopsy. ^27^ Better understanding of the genetic underpinnings of NM may allow for making the diagnosis of NM without the need for biopsy. ^32^

NM patients require multifaceted care that addresses specific needs and prevention involving wide expertise from specialists and healthcare professions. The varied presentations of NM in some patients in this review bring to light several considerations for clinical practice.

Respiratory function is commonly affected in NM patients, as 36% of the patients described had early respiratory difficulty and 56% had neuromuscular respiratory failure or weakness. Some patients experienced recurrent respiratory infection, which is cause for concern given that respiratory insufficiency, cardiopulmonary arrest, infection, and sepsis were the majority of the reported causes of death across case reports. NM patients may also experience complications post-procedures such as pleurodesis, where pneumothorax can be treated using biphasic cuirass ventilation. ^68^ Monitoring sleep and pulmonary function over time is important to assess for nocturnal hypoventilation, obstructive sleep apnea, and restrictive lung disease. One patient underwent inspiratory muscle strength training for two weeks after major surgery. This short-term intervention increased her mean inspiratory pressure and allowed her to breathe unassisted for 11-13 hours per day, compared to her medical history of restrictive lung disease and recurrent ventilatory failure post-operation. ^69^

NM has variable impacts on the motor function of each patient, and, therefore, care involves a combination of orthopedic, physical therapy, occupational/speech therapy, and neuropharmacologic agents. Speech therapy involving oral motor exercises, tongue strengthening exercises, and diadochokinesia exercises were used by one study team to help a patient with severe dysarthria improve the intelligibility of her speech. ^70^ Two case reports present rare instances where there is a role for neuropharmacologic agents in improving the life of patients with NM. The report by Sahin *et al*. documented decreased drooling and spontaneous extremity movement from a child that was previously immobile after treatment with L-tyrosine. ^71^ For a child with a *KLHL40* mutation presenting with myasthenic symptoms without antibodies directed against the acetylcholine receptor, use of the acetylcholinesterase inhibitor pyridostigmine greatly improved endurance and strength which regressed when the medication was no longer administered. ^72^

Given that the histological appearance of NM resembles that of core-rod myopathy, it is recommended that anesthesia be approached with caution in NM patients as to mitigate any theoretical risk for malignant hyperthermia particularly if a mutation in *RYR1* is present. ^73^ In a case of cleft palate repair, the team was able to avoid the use of succinylcholine by using propofol and fentanyl for induction, intubation, and adjustment of IV access, with rocuronium for facial muscle relaxation. ^74^ Intubation itself can pose challenges due to the facial dysmorphias of some patients. ^75^

For children, follow-up care should occur at regular intervals, which should be more frequent (3-4 months) for infants as opposed to older children (6-12 months). The majority of the deceased patients reviewed passed away during the first year of life, suggesting that the timing of visits for children with early onset NM symptoms may need to occur more frequently.

## Discussion

NM is a myopathy that can range from severe to mild with weakness manifesting at birth. Defects of sarcomere components or its regulators result in actin aggregates within the muscle, seen easily on electron microscopy of muscle biopsy. Several important knowledge gaps remain, including how the disease affects children across all ages. The articles surveyed in this review deepen the understanding of pediatric NM patients and provide clues that may aid in identifying subtle cases after careful evaluation.

The continued improvement in sequencing and its increased accessibility makes the possibility of identifying causative genes more feasible, including for those that produce larger proteins such as nebulin. Sequencing results confirm the heterogeneity of genes that cause NM, and several new variants were identified in pediatric NM patients which expand our understanding of how it is inherited. The majority of patients reviewed had mutations in *NEB* and *ACTA1* as has been previously reported. New examples of gonadal mosaicism leading to inheritance were identified in *ACTA1* patients by using more sophisticated sequencing modalities. Patients from non-Amish populations were found to harbor *TNNT1* mutations different from the Amish founder mutation, which resulted in a different phenotypic picture with failure to thrive and spinal rigidity rather than only progressive muscle weakness, contractures and tremors. Many of the case reports implicated nonsense-mediated decay as the mechanism by which translation of faulty transcript of NM genes is reduced, in particular for recessive form of the disease.

The cases described not only the expected presence of nemaline rods on histology, but also details about fiber size variation, filaments surrounding the rods, and inflammatory cell infiltration. These subtleties require further study to identify if these findings correlate with genotype or clinical severity.

The clinical features presented mirror many of the existing descriptions of NM, yet they highlight additional opportunities for clinical intervention to improve the lives of NM patients. A number of cases listed decreased fetal movements or antenatal ultrasound findings such as polyhydramnios as early signs; while not specific enough to be diagnostic, these may serve to document the need for a neuromuscular exam at birth. Subtle findings at birth such as hyperextension of the neck should also lead the clinician to consider a more thorough neurological examination. ^54^ Given that early respiratory difficulty was cited in over one-third of patients, more needs to be done to identify ways to provide respiratory support for neonates with suspected NM with special attention to the various craniofacial deformities found in some patients. Respiratory health is critical, considering the number of patients described as experiencing recurrent infection and dying from pulmonary complications particularly in the first year of life.

Little was mentioned about management of spinal curvature despite almost 40% of patients experiencing this complication. Amongst these cases, there were examples of both dilated and hypertrophic cardiomyopathy in patients with mutations with mutations in *ACTA1, TNNT1*, and *MYO18B*. Cardiomyopathy had been reported in *ACTA1* and Amish *TNNT1* patients previously, however this report of a pediatric patient with a mutation *MYO18B* was one of the first that showed an association with hypertrophy. ^17^ The case of *MYO18B* also highlight that cardiac phenotypes seen in model organisms may serve to identify genes that may later be found to be important in skeletal muscle and be linked to NM pathology. The paper by Saito *et al*. introduces the point that normal intelligence may not be common to all patients with NM and therefore emphasizes the need for more detailed analysis of the pediatric brain. ^55^ Lastly, these cases collectively underscore how presentation is not consistent across patients with mutations in the same gene; future studies examining patients with different mutations in the same gene should carefully track the precise symptoms that may distinguish severity.

To compile the most recent clinical reports of NM, this review is limited by the findings shared in the literature. Therefore, the trends identified in the results are based on the information explicitly described in the case reports, which varied in their detail (Table 2). The lack of histological or genetic results for some patients reduces the correlations that can be made between genotype and phenotype. Studies that include a large cohort of patients with NM with different genetic mutations are needed to identify such patterns, especially if sequencing and testing are done in a comparable manner. These studies will allow for more robust correlation between genotype and phenotype that may be useful in predicting prognosis.

Greater interplay between clinical and basic science findings will be critical in developing clinical prevention and treatment strategies. Future studies may consider tests, procedures, and treatments for NM. For example, further study should examine the value of imaging, as suggested by Ennis et al. ^27^ The value of L-tyrosine as treatment should also be considered, although there have been mixed results in human and animal studies. Ryan et al. describe improvement in bulbar function, activity level and exercise tolerance in four patients who received L-tyrosine supplementation; however, the underlying genetic change was only identified in one patient in *TPM3*. ^77^ A study in mice with a mutation in *ACTA1* showed similar improvements in improving mobility in addition to showing a decrease in rods and degenerating fibers on muscle biopsy. ^78^ However, work in zebrafish models of nebulin NM have not shown these changes after administration of L-tyrosine or other supplements. ^79^ These findings are similar to a study using two established mouse models and one zebrafish model with *ACTA1* mutations. ^76^ Continued genetic studies are needed to characterize the responsible genes that cause other forms of NM. More reports sharing best practices in improving quality of life for NM patients would benefit the field, particularly related to respiratory and orthopedic care that would benefit many of the patients reviewed.

Overall, this review demonstrates the progress that has been, and is yet to be, made in the diagnosis and mechanistic understanding of NM in children. More research, particularly into the importance of sarcomeric structures and regulatory proteins in maintaining healthy muscle, will be needed. These concepts will be critical to establishing best practices and standards for diagnosis and treatment in pediatric NM.

## Supporting information

PRISMA checklist

## Data Availability

All data produced in the present work are contained in the manuscript.

